# Integrative genetic analysis reveals new relationships between intraocular pressure, glaucoma and ischemic stroke risk: A study based on combined SNP-to-gene, mendelian randomization and pathway investigations

**DOI:** 10.1101/2024.08.26.24312564

**Authors:** Qi Zhang, Di Hu, Zenan Lin, Junhong Jiang

## Abstract

**Background:** Ischemic stroke (IS) is a leading cause of death in elderly people. Previous studies on exploring the association between intraocular pressure (IOP), glaucoma and the IS risk provided inconsistent results and unclear elucidations.

**Methods:** Here, multiple genetic approaches were employed to investigate the possible causality between these traits. First, we performed the traditional bidirectional mendelian randomization (MR) study to explore the causal relationship between IOP, glaucoma and IS. Second, the SNPs selected as instrumental variables for IOP and primary open-angle glaucoma (POAG) were mapped to relevant genes by the novel combined SNP-to-gene (cS2G) method. The genes with significant causal effects on IS were then introduced to the gene ontology (GO), pathway and colocalization analyses. Third, the partitioned heritability analysis was also performed to evaluate the genome complexity with the Linkage Disequilibrium Score (LDSC) tool. Fourth, we also performed single SNP mendelian randomization (SSMR) study to find the SNPs of IOP and glaucoma, which had significant causal influence on IS risk. Then, they were introduced to the cytogenetic investigation. The multiple variable MR (MVMR) was applied to assess the independence of the causal effect of the exposures.

**Results:** The MR results supported the view that the elevated IOP and POAG may contribute to the IS risk, but not vice versa. By using the cS2G approach, we identified 31 and 3 genes which may play key roles in the IOP- and POAG-induced IS risk, respectively. The GO and pathway analyses indicated the olfactory pathway to be a crucial pathway in the IOP-associated mechanism. The colocalization study strengthened the causal implications of genes *CDKNA2A* and *CDKN2B-AS1* between POAG and IS. The partitioned heritability analysis showed that the most enriched categories for both IOP and POAG were regulatory-associated terms such as the Super-enhancer. The SSMR study demonstrated that the IOP-associated SNPs with causal effects on IS were located majorly in chromosomes 1 and 11, while the POAG-associated ones were mostly found in chromosomes 9 and 4. The MVMR confirmed that the causal effects of IOP and POAG were not independent from each other.

**Conclusions:** This work provides novel evidences to support the causal implications between IOP, POAG and IS, and offered putative pathway and genes for managing IOP and POAG in IS.

## 1. Introduction

Ischemic stroke (IS) remains a significant global health concern, representing one of the major causes of disability and mortality ^1^. The pathogenesis of ischemic stroke involves the occlusion of cerebral blood vessels, resulting in the shortage of blood supply to affected brain areas and subsequent regional damage. While hypertension, diabetes mellitus, and smoking are acknowledged to contribute to stroke risk, emerging evidence suggest that factors beyond the conventional cardiovascular spectrum may also play a role in stroke pathophysiology ^2, 3^.

Recently, there has been growing interest in studying the potential association between intraocular pressure (IOP) or glaucoma, and IS ^4, 5^. IOP, a physical measure of the fluid pressure within the eye, is primarily regulated by the balance between aqueous humor production and outflow. Though the elevated IOP may not be detected in the normal tension glaucoma, it is a hallmark feature of other glaucoma types, and believed to be the leading cause for the progressive retinal ganglion cell loss and visual field defects ^6^. While the primary pathological consequence of elevated IOP is optic nerve damage and visual impairment, recent works have also indicated that elevated IOP may also be associated with systemic vascular dysfunction, including the alterations in cerebral blood flow dynamics ^7^.

In detail, the rationale for investigating the relationship between IOP and IS stems from the following aspects. First, both elevated IOP and IS share common risk factors, such as hypertension, diabetes mellitus, and metabolic syndrome ^2, 8–10^. Second, previous investigations have reported the anatomical and physiological connections between the ocular and cerebrovascular systems, indicating that alterations in IOP may contribute to the stroke risk through impacting the intracranial pressure ^11^. Third, previous population-based studies and clinical cohorts has provided preliminary insights into the plausible link between elevated IOP and increased stroke risk, although findings have been contradictory and warrant more in-depth investigations ^12^. Given the potential implications for stroke prevention and management, clarifying the relationship between IOP and ischemic stroke seems to be a promising research topic. A better understanding of the underlying mechanisms linking ocular and cerebrovascular health may provide novel insights into stroke pathophysiology and offer new therapeutic targets for stroke prevention and management.

Mendelian Randomization (MR) analysis is a powerful biomedical method that uses appropriate single nucleotide polymorphisms (SNPs) as instrumental variables (IVs) to identify the potential causal effects between the exposure and outcome traits. Since the conventional 2-sample MR was not able to provide the detailed portrait of the underlying mechanism, an up-to-date combined SNP-to-gene (cS2G) strategy^13^ was employed to demonstrate the key genes and pathways involved in the causality. By leveraging multiple genetic tools, herein, we investigated the causality between glaucoma, IOP and ischemic stroke, and further identified relevant genes and pathways for future biomarker research.

## 2. Methods

### 2.1 GWAS data source of intraocular pressure, glaucoma and ischemic stroke

Inn this work, numerous GWAS datasets for intraocular pressure, glaucoma and ischemic stroke were introduced for further analyses. The IOP (both of right and left eye) were assessed by the gold standard Goldmann method. Four glaucoma-associated datasets, i.e. primary open angle glaucoma (POAG), primary angle closure glaucoma (PACG), normal tension glaucoma (NTG) and undefined glaucoma) were also analysed in this study. Besides, a GWAS dataset on analysing IS was selected for further investigations (Malik, Chauhan et al. 2018). The GWAS summary statistics data of these datasets were obtained from the IEU OpenGWAS project (https://gwas.mrcieu.ac.uk/). Their basic features (GWAS ID, sample size, ethnics and links) were summarized in Table S1.

### 2.2 Selection of genetic instrumental variables (IVs)

The summary statistics data of IOP contained 9851867 single nucleotide polymorphisms (SNPs) data. 85 SNPs with P values less than 5E-8 and valid Reference SNP identifications (rsIDs) were selected as significant SNPs (See supplementary Table S2.) By using R package TwoSampleMR (version 0.5.6) [31], we performed the linkage disequilibrium (LD) to acquire independent SNPs (r2 < 0.001 within 10Mb range). We set the parameter ‘pop’ to ‘EUR’ to use 1000 Genomes Project Phase 3 (EUR) as the reference panel. The absolute value of five F statistics (see Supplementary Table S2.) were all larger than 10 indicating sufficient statistical strength of the selected IVs [33]. The selection process of IVs for IS and various kinds of glaucoma subtypes were similar to the abovementioned procedure. Their IVs were summarized in Table S2-S7.

### 2.3 The univariable, bidirectional MR and multiple variable MR (MVMR)

The exposure and outcome datasets were harmonized. The proxy SNPs (R2 > 0.8) were obtained when no SNP was found in the outcome dataset. Then, five MR analytical methods (IVW (Inverse variance weighted, Weighted median, Weighted mode, MR Egger, Simple mode)) were employed to provide a comprehensive MR results. Each trait was introduced to the MR analysis as exposure and outcome, respectively. Besides, the MVMR analyses were performed on the traits which were found to have causal influence on the IS risk in the univariable MR investigations.

### 2.4 The Linkage disequilibrium score regression (LDSC) and partitioned heritability analysis

In the current work, the IOP of the right eye was selected for the MR study. However, the eyes are symmetric (namely right and left eye) organs, thus, there is also a GWAS dataset of the IOP of left eye. In order to prove that there is a strong correlation between the IOP of right and left eye, we performed a genetic correlation analysis between them with LDSC tool. The result indicates a strong genetic correlation (*P*= 3.9755e-86) between the IOP of right and left eye (See supplementary Table S8), which guarantees the sufficiency of the utilization of the GWAS data of right eye.

In the SNP partitioned heritability study, 96 previously developed baseline genomic molecular annotations (See Table S9; 1000G_Phase3_baselineLD_v2.2_ldscores were downloaded from https://doi.org/10.5281/zenodo.10515792) such as promoter, super-enhancers, conserved regions and etc ^14^. SNP-h2 partitioning implemented in the LDSC approach allowed us to evaluate simultaneously the functional annotations, and estimate their enrichments.

### 2.5 The cS2G, Gene ontology (GO) and pathway analyses

Under the guidance of the novel cS2G method ^15^, the genes correlated to the critical SNPs for IOP and glaucoma were identified. Then, they were introduced to the MR independently and those lead to significant MR causal effect on IS risk were selected for further colocalization, GO and pathway analyses.

### 2.6 Colocalization analysis

The genes of interest were introduced to the colocalization analysis. In detail, we performed the colocalization study in the specific region of each gene and tested whether identified relation between two phenotypical traits were driven by linkage disequilibrium within the given locus. Theoretically, the approximate Bayes Factor colocalization analyses was conducted under the following five hypotheses, 1. neither trait had an association in the given locus; 2. only trait 1 had an association in the locus; 3. only trait 2 had an association in the locus; 4. both traits had associations, but with different causal variants; 5. both traits had associations and shared one identical causal variant. In the end, five posterior probabilities (PH0, PH1, PH2, PH3, and PH4) were generated for each hypothesis ^16^.

### 2.7 Cytogenetic analysis

Using the SSMR (single SNP mendelian randomization), the single SNP was introduced to the MR separately, and those with a *P* value less than 0.05 were identified to be significant SNPs. Their cytogenetic information was investigated.

### 2.8 Statistical analysis

Using R packages TwoSampleMR (version 0.6.6),ieugwasr (1.0.1), gwasglue (0.0.0.9000), we performed the following analyses. The selected IVs’ summary statistics were harmonized with outcome GWAS dataset. The IVW method was regarded as the major analytical tool. As complementary sensitivity tests, the weighted median, weighted mode, simple mode, the MR-Egger regression, and leave-one-out methods were introduced to assess the heterogeneity and pleiotropy, and evaluate the robustness of identified causal associations. R packages coloc (5.2.3) and locuscomparer (1.0.0) were employed to conduct the colocalization analysis. The R package qqman (0.1.9) was used to demonstrate the mannhattan plot. The clusterProfiler (4.12.0) package was sued to perform the GO and pathway analysis. For the LDSC analysis, the LDSC (LD SCore v1.0.1) command tool was utilized under the Python (3.12.4) environment. A *P* value less than 0.05 was regarded as statistically significant.

## 3. Results

### 3.1 The IOP and POAG have causal effects on the IS risk

As shown in Figure 1., multiple genetic analytical approaches were employed to explore the associations between IOP, glaucoma and IS. In order to clarify the potential causality, each trait was investigated as exposure as well as outcome. However, no SNP in the PACG summary statistics had a significant *P* value less than 5E-8, it was analysed only as outcome in the current work. Standing in line with previously acknowledged view, the increase of IOP has an obvious causal effect on the risks of various kinds of glaucoma subtypes (i.e. primary open-angle glaucoma(POAG), primary angle-closure glaucoma(PACG), normal tension glaucoma (NTG)) (See Figure 2. and supplementary Table S10). Despite NTG, our result showed that the genetically proxied glaucoma, PACG and POAG also had causal influence on the IOP. (See Figure 2. and Table S11).

**Figure 1.**
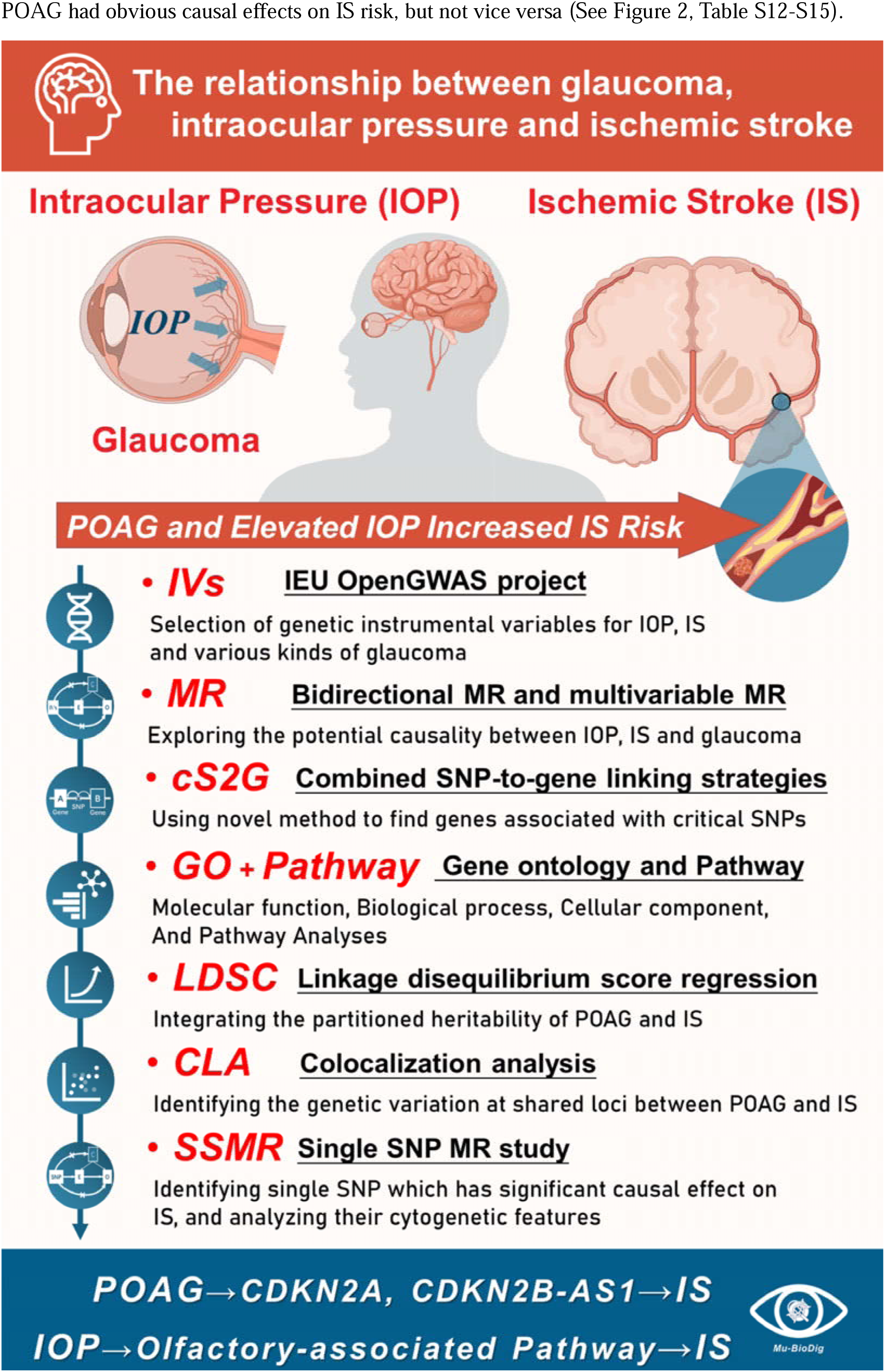
The workflow for the study. Multiple genetic analytical tools, such as MR, cS2G, GO, colocalization, LDSC were employed to evaluate the associations between IOP, glaucoma and IS.

**Figure 2.**
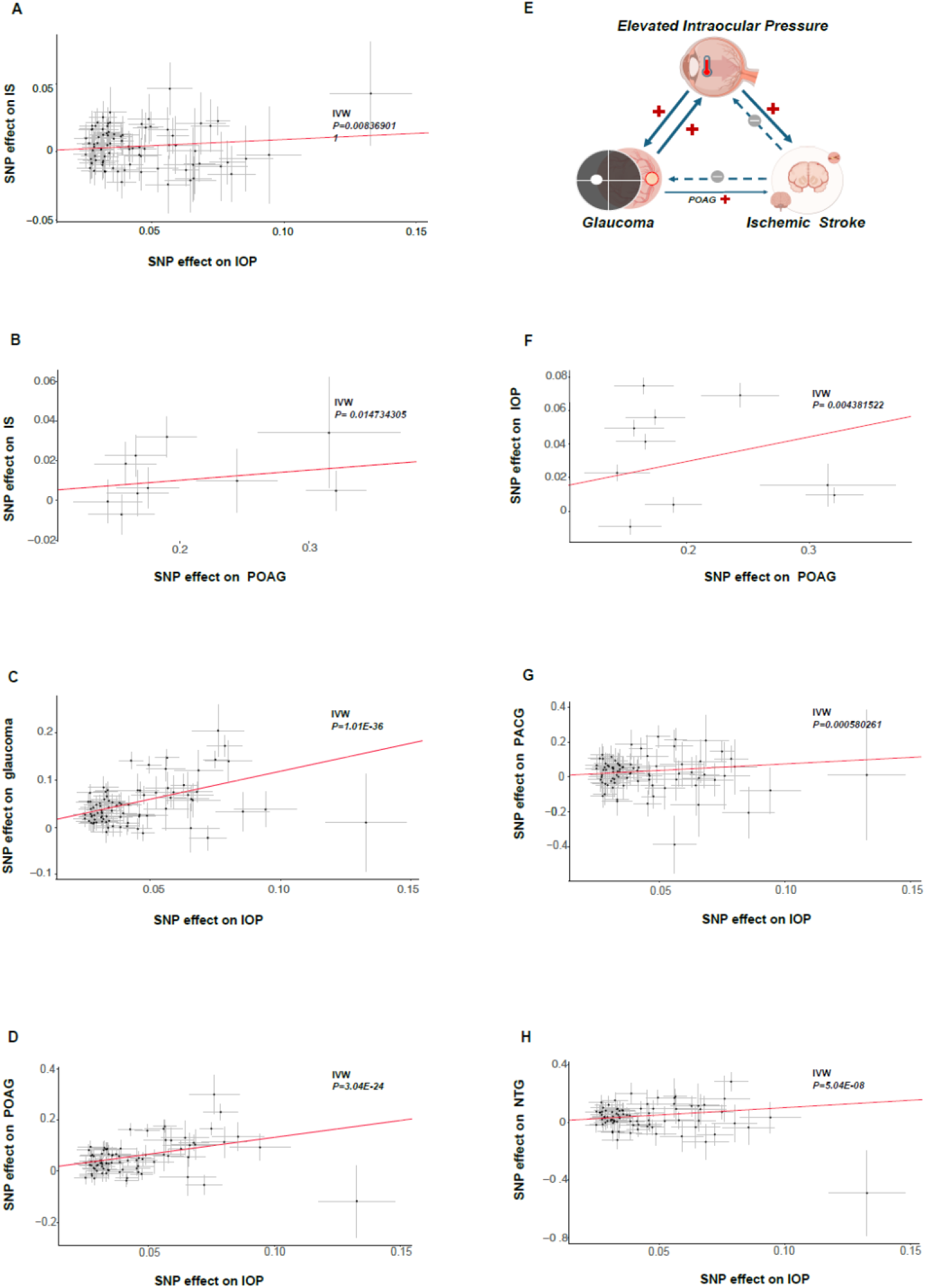
The bidirectional MR conducted on IOP, various kinds of glaucoma types and IS. Scatter plots for MR analyses of the causal influence of IOP on IS (A), POAG on IS (B), IOP on glaucoma (C), IOP on POAG (D), POAG on IOP (F), IOP on PACG (G), IOP on NTG (H). The schema demonstrating the results of the bidirectional MR (E).

We detected no remarkable causal relationship between glaucoma, NTG, PACG and IS (See Figure 2, Table S12 and S13). Notably, our results indicated that the genetically proxied IOP and POAG had obvious causal effects on IS risk, but not vice versa (See Figure 2, Table S12-S15).

### 3.2 Using novel combined SNP-to-gene (cS2G) approach to identify significant genes

As the MR study suggested that the genetically predicted IOP and POAG had causal effects on IS risk, they were introduced into the further cS2G analyses. First, the SNPs with a *P* less than 5E-8 were selected (See Figure 3.A). Their corresponding genes were identified by searching the archive of the cS2G dataset. In order to obtain a relatively complete result, the cS2G datasets generated on two cohorts (i.e. UK Biobank and 1000 EUR genome) were simultaneously investigated. Finally, the corresponding genes from UK Biobank and 100 EUR genome were aggregated. By using this cS2G method, we identify 395 genes from 2406 SNPs of the IOP GWAS dataset. (see Table S16.) Among them, 378 were successfully introduced to the single gene MR on IS. The result indicated 31 genes to have significant causal effect on IS (*P* < 0.05). (See Table S17.) They were introduced into the Gene Ontology (cellular component, molecular function and biological process) and pathway analyses, and the result indicated the olfactory pathway to play a key role in the IOP-induced IS risk. Similarly, the same approach was introduced to study POAG. Three critical genes (i.e. *CDKN2A*, *CDKN2B-AS1* and *GAS7*) were found to potentially participate in the POAG-induced IS risk (See Table S18-S19).

**Figure 3.**
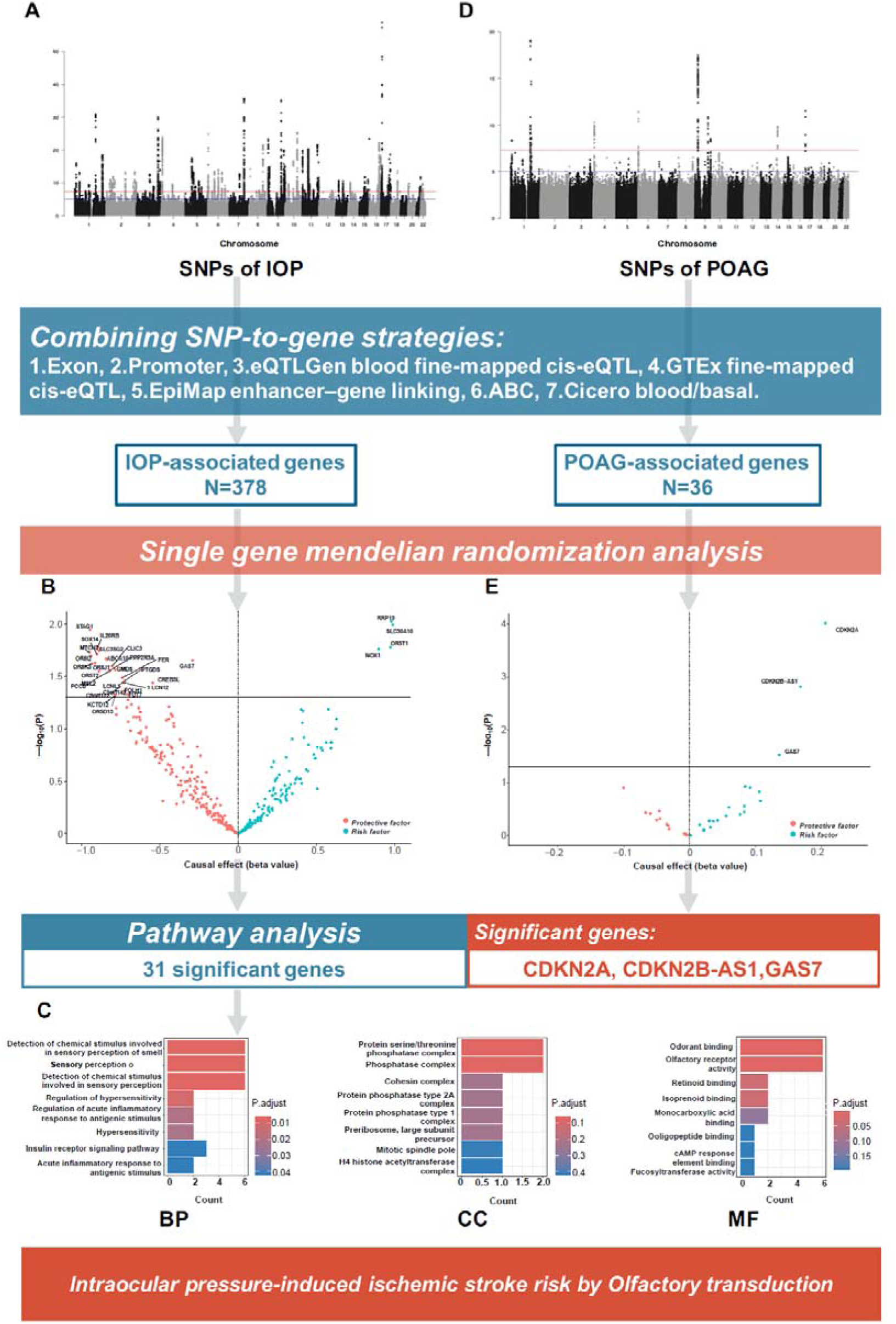
A and D) The Manhattan plot demonstrating significant SNPs of IOP and POAG. B and E) By using the cS2G method, 378 and 36 genes were identified to be the IOP-and POAG-associated genes, respectively. The volcano plots highlighting the genes with significant causal effects on IS. C) The single gene MR analysis indicated *CDKN2A*, *CDKN2B-AS1* and *GAS7* to play key roles in the POAG-induced IS risk. The GO and pathway analyses with 31 IOP-associated genes indicated the olfactory pathway may contribute to the IOP-induced IS risk.

### 3.3 The Approximate Bayesian colocalization analyses

As shown in Figure 4., we gained strong evidences to support the 3rd Hypothesis for both *CDKN2A* (posterior probability = 0.92) and *CDKN2B-AS1* (posterior probability = 0.92), which means that POAG and IS had associations in the given locus of these two genes, however, with different causal variant. The study on *GAS7* demonstrated that only POAG had associations in the given region.

**Figure 4.**
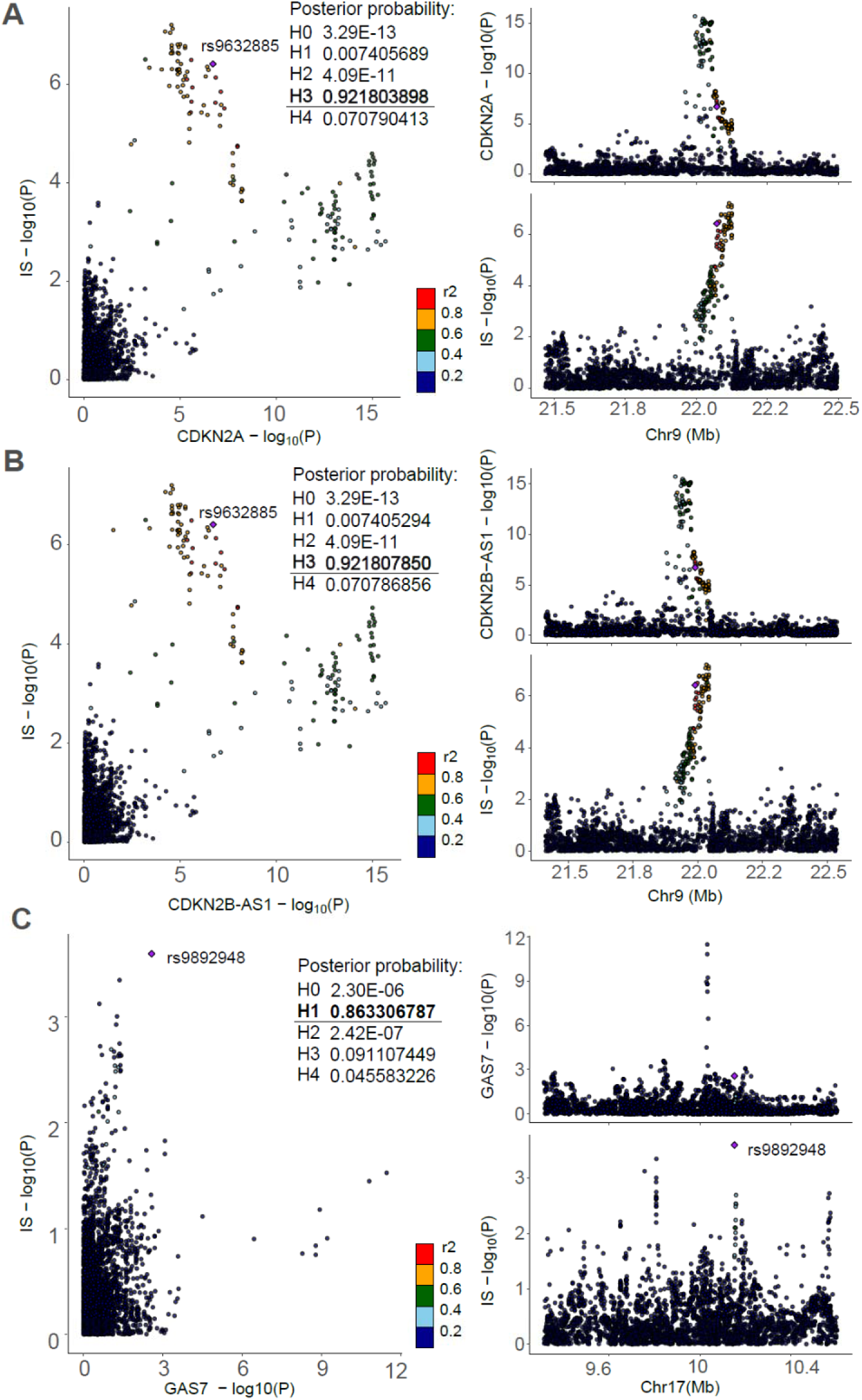
The colocalization analyses on the POAG-associated genes *CDKN1A* (A), *CDKN2B-AS1* (B) and *GAS7* (C) and IS. The results demonstrated that both *CDKN2A* and *CDKN2B-AS1* had strong evidences (Posterior probability>0.8) to support the 3rd Hypothesis. However, *GAS7* lacked the evidence to support neither the 3rd nor 4th Hypothesis.

### 3.4 Partitioned heritability by multiple functional categories

In order to accurately articulate the complexity of the genome of POAG and IS, we partitioned their heritability into 96 functional categories, by using the baseline model provided by Finucane et. al.^17^. Intriguingly, both traits had statistically significant (less than Bonferoni-corrected *P*, i.e. 0.05/96) enrichments in three categories. In detail, SuperEnhancer_HniszL2_0 (Enrichment = 2.923, *P* = 0.0001), H3K27ac_HniszL2_0 (Enrichment =2.546, *P* = 1.51E-5) and H3K27ac_PGC2L2_0 (Enrichment =4.288, *P* = 0.0001) were identified to be significant in POAG. Of notice, the SuperEnhancer_HniszL2_0 (Enrichment = 2.186, *P* = 1.26E-6) was also detected in ischemic stroke (See Figure 5. and Table S20-S21.).

**Figure 5.**
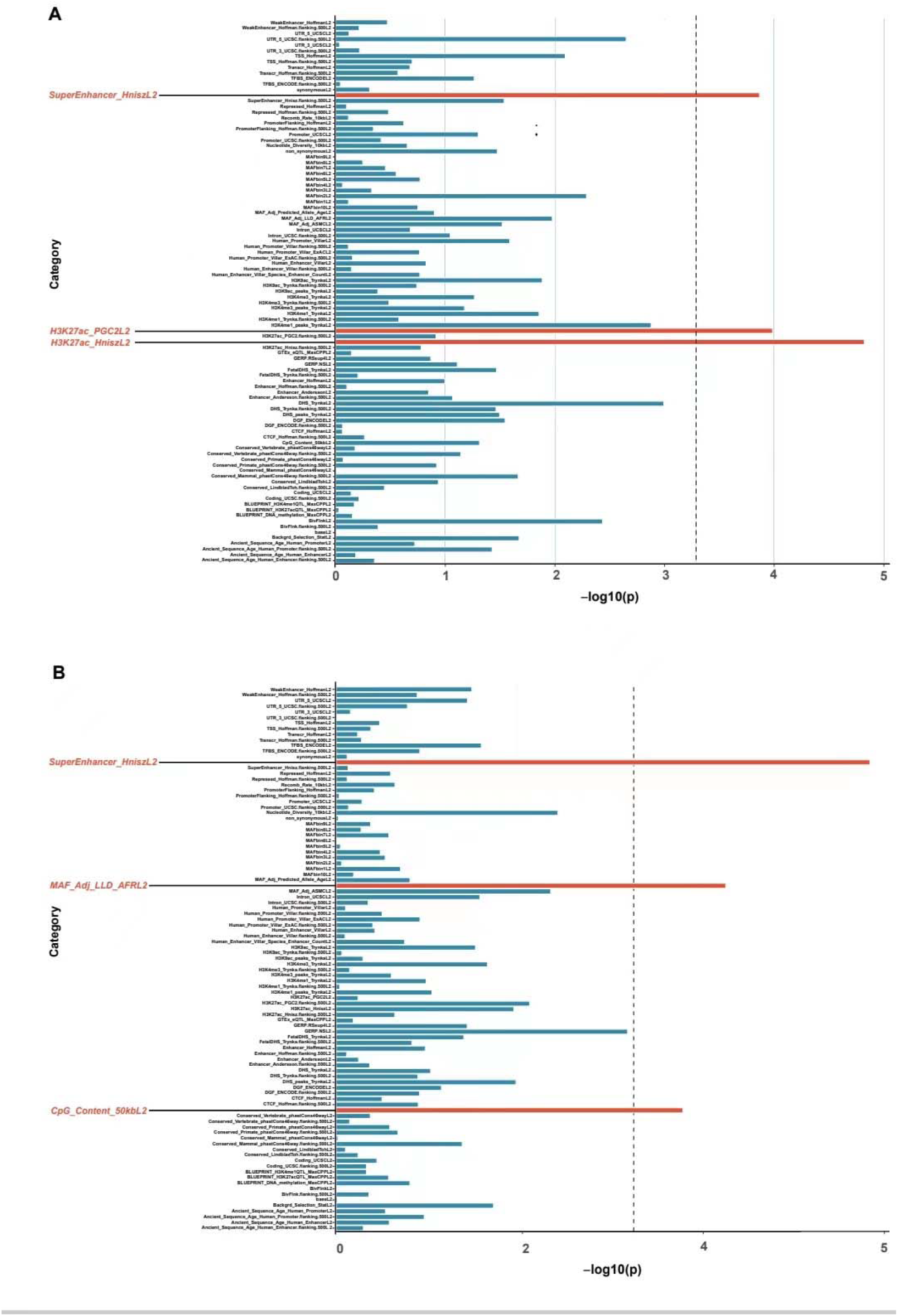
The partitioned heritability analyses on POAG (A) and IS (B).

### 3.5. Single SNP mendelian randomization (SSMR)

SSMR revealed 278 and 507 IOP-associated variants with significant (*P* < 0.05) positive and negative causal estimates on IS, respectively (Figure 6A-B and Table S22). More than one third of the positive causal SNPs were on chromosomes 1, whereas over 50% of negative causal SNPs were on chromosomes 11.

**Figure 6.**
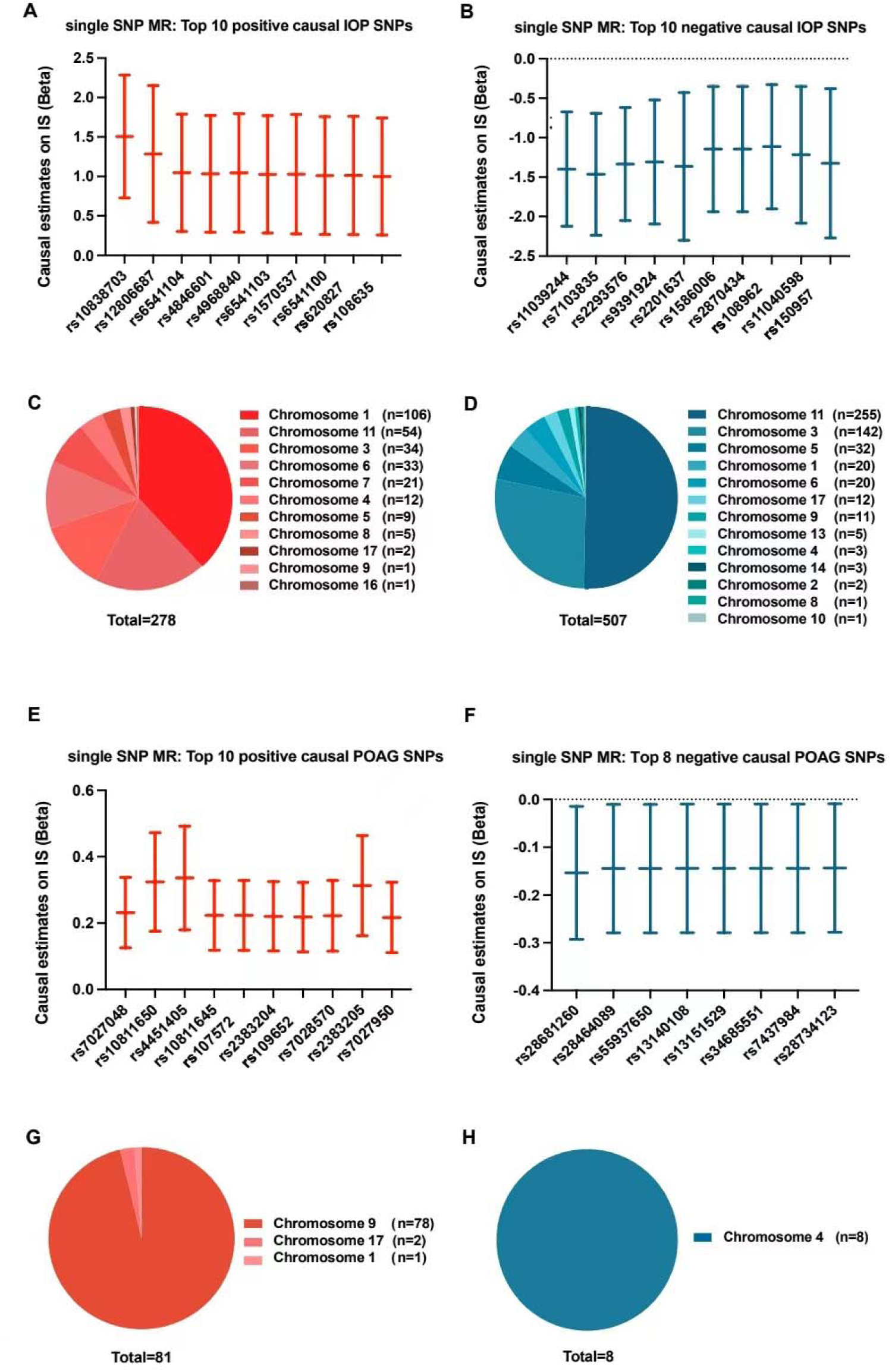
The cytogenetic analyses of SNPs with significant causal effects on IS. (A and B) The plots demonstrating the top 10 IOP-associated SNPs with positive and negative causal influences on IS risk, respectively. (C and D) The pie charts showed the chromosome distribution of the significant IOP-associated SNPs with positive and negative causal estimates. (E and F) The plots demonstrating the top POAG-associated SNPs with positive and negative causal influences on IS risk, respectively. (G and H) The pie charts showed the chromosome distribution of the significant POAG-associated SNPs with positive and negative causal estimates.

In comparison to IOP, we found less POAG-associated variants with significant positive (n=81) and negative (n=8) causal estimates on IS. (Figure 6.E-F and Table S23). The positive causal SNPs were majorly located on chromosomes 9, and all of negative causal SNPs were on chromosomes 4.

### 3.6 MVMR

By performing the MVMR, we tested whether the POAG’ causal influence on IS was independent from IOP. Though both traits were proved to have significant causal effects on IS risk in the univariable MR, neither POAG nor IOP demonstrated remarkable influence in the MVMR (See Figure 7. and Table 24).

**Figure 7.**
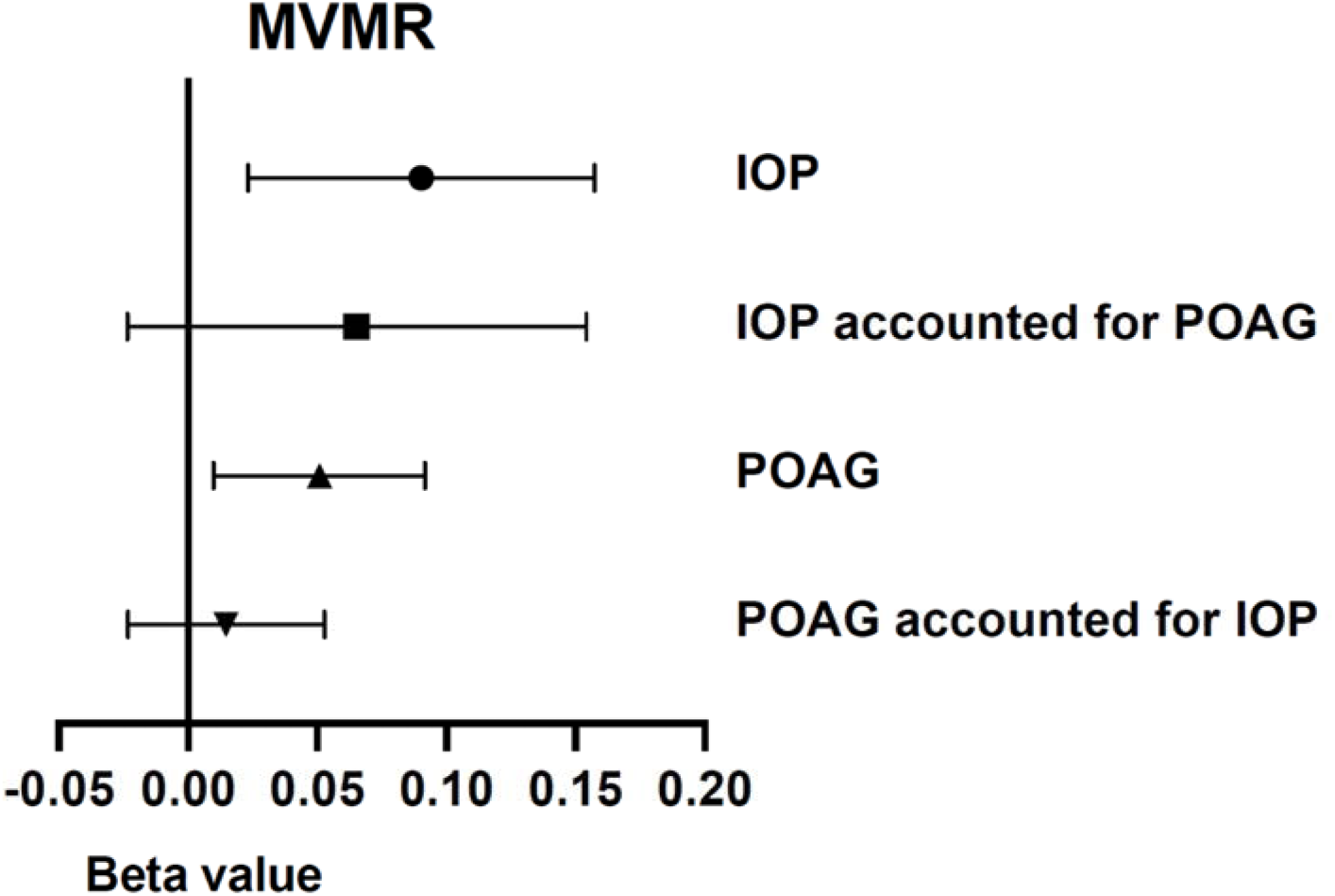
The forest plot of MVMR demonstrate that the causal effects on IS of IOP and POAG were not independent from each other.

## 4. Discussions

Ischemic stroke (IS) is a leading cause of disability and represents a major and growing public health problem worldwide. In this work, our integrative genetic analysis based on genome-wide association studies (GWAS) datasets revealed a significant positive genetic correlation of POAG with IS and confirmed for the first time there is the common genetic variation at both genome and transcriptome levels between the two disorders. Additionally, we further discovered that elevated IOP, a primary feature of glaucoma, may increase the risk of IS by the potential mechanistic link mediated through the olfactory pathway. The findings of this study provide compelling evidence to support a causal relationship between IOP, POAG and IS, which provide the novel insights into development of potential therapeutic targets for stroke prevention and treatment.

Glaucoma, as a potential risk factor for IS, has attracted increasing attention from researchers and clinicians. The correlation between glaucoma and stroke has been observed in previous observational studies. Ho and colleagues conducted a population-based, prospective cohort study with a large sample size of 4032 POAG patients and 20160 control participants from the Taiwan National Health Insurance Research Database, they found that 1.5-fold increased risk of stroke in POAG patients compared with population controls, when followed for 5 years^18^. A similar result in the US veteran population was observed by French et al.^19^, who found that the risk of stroke was higher in patients with POAG than in controls (6.4% vs 4.8%). Likewise, a recent meta-analysis involving 362267 participants showed an association of POAG with a higher risk of stroke^5^. Consistent with previous studies, our genetic analysis results show that POAG had obvious causal effects on IS risk.

While several studies found a correlation between POAG and IS, the exact mechanism and causal direction behind these observations have not yet been investigated clearly. In the present study, further analysis by co-localization showed that the variants in *CDKN1A* and *CDKN2B-AS1* two genes play key roles in the pathogenesis of POAG and IS. The cyclin-dependent kinase inhibitor 2A (*CDKN2A*) gene and the cyclin-dependent kinase inhibitor 2B antisense noncoding RNA (*CDKN2B-AS1*) gene are both located on chromosome 9p21, a region that has been found to be associated closely with a wide range of human diseases, including cancer^20^, cardiovascular disease^21, 22^ and neurodegenerative diseases^23^. *CDKN2A* gene encodes p16 and p14, two crucial cell cycle regulators, involved in cell cycle progression^24^, differentiation^25^, senescence^26^, and apoptosis^27^. While the *CDKN2B-AS1* gene, also known as *ANRIL*, transcribes into a long noncoding RNA in the antisense direction that is involved in modulating the nearby *CDKN2A/2B* genes by interacting with polycomb proteins, and subsequently participating in the alteration of cell cycle regulation^28, 29^. To date, the *CDKN2A* and *CDKN2B-AS1* genes has been reported to be associated with IS. Several human genetic research suggested that the *CDKN2A* gene polymorphism is associated with IS in different populations including Northern European^30^, Chinese^31^ and African Americans^32^ and West Africans^33^. Similarly, the genetic susceptibility of *CDKN2B-AS1* has been shown to be closely related to the occurrence and outcome of IS.^34–36^ On the other hand, *CDKN2A* and *CDKN2B-AS1* genes were also reported as the significantly mutated gene in the pathogenesis of POAG.^29, 37, 38^

Basic research also provided evidence for the potential roles of *CDKN2A* and *CDKN2B-AS1* in the pathogenesis of POAG and IS. In the animal models of glaucoma, multiple *CDKN2B-AS1* splice variants are observed in rat retina, furthermore, CDKN2A protein expression is significantly upregulated in rat retina, and this elevated expression corresponds to ongoing RGC death.^29^ *CDKN2A* encodes crucial cell cycle regulators that affect cell proliferation or senescence in RGCs by regulating cell cycle regulation, thereby contributing to POAG pathogenesis.^39, 40^ Mice with their *CDKN2B-AS*1 gene partially deleted show increased RGC vulnerability to apoptosis.^40^ Meanwhile, *CDKN2A* and *CDKN2B-AS1* were also shown to the regulate vascular smooth muscle cell proliferation^28^, and the accumulation of vascular smooth muscle cells can contribute to the vessel stenosis^41^, which in turn leads to an increased risk of IS.^42^ In the mice model of IS, the level of CDKN2B-AS1 exhibited a significant increase in the brain, and downregulation of CDKN2B-AS1 can reduce neuroinflammation by negatively regulating miR-671-5p to inhibit NF-κB.^43^ The latest GWAS results for POAG loci showed that *CDKN2A* and *CDKN2B-AS1* might be associated with vascular traits.^37^ These evidences may, partly, account for the genetic association between POAG and IS. A recent MR study found a positive causal effect value (OR=1.03) in the POAG-ischemic stroke association, however, the *P* value was above the 0.05 threshold.^44^ This might be owed to their relatively small sample size (n=63412, in comparison to the current dataset with a sample size of 214634 participants) of the POAG GWAS dataset, which may limit the statistical power of MR analysis.

Leveraging genome-wide association studies (GWAS) analysis, we identified significant genetic variants associated with both elevated IOP and increased risk of IS, supporting the notion of a shared genetic basis between these two phenotypes.^45^ Our results corroborate previous epidemiological studies suggesting an association between elevated IOP and ischemic stroke risk.^46, 47^ While the precise mechanisms underlying this association have remained elusive, our findings offer novel insights into the potential involvement of the olfactory pathway in mediating the observed relationship. Specifically, we identified genetic variants implicated in olfactory receptor activity and olfactory transduction pathways that were significantly associated with both IOP and ischemic stroke risk.^48–52^ This provides compelling evidence suggesting that alterations in olfactory function may represent a mechanistic link between elevated IOP and increased stroke risk.

The olfactory pathway serves as a critical interface between the external environment and the central nervous system, playing a fundamental role in odor detection and processing. Recent studies have highlighted the olfactory system’s role beyond sensory perception, implicating it in various physiological and pathological processes, including neurodegenerative diseases, cognitive decline, and cardiovascular health.^53–58^ Our findings extend this understanding by implicating olfactory dysfunction as a potential mediator linking elevated IOP to ischemic stroke risk. Several potential mechanisms may underlie the observed association between olfactory dysfunction and stroke risk. Firstly, olfactory dysfunction may serve as a marker of systemic vascular dysfunction, reflecting shared pathophysiological processes underlying both conditions.^56, 59^ Secondly, alterations in olfactory receptor activity and olfactory signaling pathways may directly impact cerebrovascular health through mechanisms such as inflammation, oxidative stress, and endothelial dysfunction.^60–63^ Finally, disruption of olfactory pathways may influence autonomic nervous system function, thereby modulating cardiovascular risk factors and contributing to stroke pathogenesis.^56, 64^

While our study provides evidence supporting the causal relationship between IOP, POAG and ischemic stroke risk, several limitations should be acknowledged. First, the observational nature of GWAS analysis precludes establishing causality definitively. Further mechanistic studies using animal models and experimental manipulations are warranted to validate our findings and elucidate underlying biological mechanisms. Second, previous studies have reported a potential association between the intracranial pressure (ICP) and IOP, thus, a mediated MR may help clarify whether the elevated IOP induces the IS risk through the ICP-associated pathway ^65, 66^. However, to our knowledge, a GWAS dataset on ICP has not been released yet. Third, the findings were concluded from the datasets with European participants, the generalizability of our results to diverse populations and ethnic groups warrants further investigation.

## 5. Conclusion

In conclusion, our study provides novel insights into the relationship between intraocular pressure, POAG, and ischemic stroke risk. By leveraging GWAS analysis, we have identified genetic variants implicating the olfactory pathway as a potential mediator of the elevated IOP-induced ischemic stroke risk, and the *CDKN2A* and *CDKN2B-AS1* genes of the observed association between POAG and ischemic stroke. Elucidating the mechanisms linking ocular and cerebrovascular health may offer new avenues for stroke prevention and management, highlighting the importance of interdisciplinary approaches in understanding complex disease pathogenesis.

## Data Availability

The links of the GWAS data were described appropriately in the paper. The codes and detailed information required to replicate the results in this work are available from the corresponding authors upon reasonable request.

## Author Contributions

Conception, supervision and administration: Zenan Lin and Junhong Jiang; Data curation: Qi Zhang and Di Hu; Investigation: Qi Zhang and Di Hu; Methodology: Zenan Lin and Junhong Jiang; Writing – original draft: Qi Zhang and Di Hu; Writing – review & editing: Zenan Lin and Junhong Jiang.

## Funding

No fund was obtained for this work.

## Conflict of Interest

The authors declare no conflict of interests.

## Ethics Approval

The GWAS data were publicly available and approved by their original institutions. An ethics approval for the current work is not required.

## Acknowledgement

The authors appreciate the participants and investigators who made the GWAS datasets publicly available.

## Notes

### Competing Interest Statement

The authors have declared no competing interest.

### Funding Statement

This study did not receive any funding

### Author Declarations

The GWAS summary statistics data of these datasets were obtained from the IEU OpenGWAS project (https://gwas.mrcieu.ac.uk/). Their basic features (GWAS ID, sample size, ethnics and links) were summarized in Table S1.

